# Modeling the Competitive Evolution and Large-scale Transmission Dynamics of SARS-CoV-2 and Emerging Viral Pathogens

**DOI:** 10.1101/2025.02.15.25322128

**Authors:** Sisi Huang, Xiangyun Lu, Zhengyun Xiao, Yiqi Zhuang, Xin Wei, Zuyan Fan, Lingtong Huang, Shijie Zhao, Michael Lynch, Hangping Yao, Min Zheng, Chao Jiang

**Author notes:** These authors contributed equally to this work (Sisi Huang, Xiangyun Lu, Zhengyun Xiao). Correspondence (Chao Jiang) (Min Zheng) (Hangping Yao).

## Abstract

A fundamental question in virology and epidemiology is how viral variants compete and evolve during large-scale transmission and what drives one variant to dominate—key to understanding pathogen emergence and mutation-driven fitness advantages. In this study, we address this by integrating epidemiological, clinical, and experimental data to examine the co-infection and competitive evolution of the earliest SARS-CoV-2 lineages, A and B, two key variants circulating in early 2020. To capture these dynamics, we developed the evolutionary-SIR-Seeding-Spreading (evoSSS) model, a holistic framework that integrates transmission, competition, and spatial bottlenecks. The model simulates viral competition across multiple scales, from intra-host replication to inter-host transmission and from regional outbreaks to global spread. It also predicts risks for other infectious pathogens, such as influenza and monkeypox, and clarifies the competitive interactions between SARS-CoV-2 and influenza viruses. This study provides valuable insights into viral competitive evolution and informs strategies for managing future pandemics.

## Introduction

The rapid spread of known and unknown pathogens poses a significant global health challenge. Understanding how viral variants evolve and compete during large-scale transmission is crucial for predicting which variants will dominate and the fitness advantages conferred by specific mutations. This knowledge is fundamental to anticipating pathogen emergence and informing response strategies. However, the acquisition of comprehensive data to study these dynamics at a population scale was not feasible until the COVID-19 pandemic^1^.

Beginning in December 2019, the unprecedented global genomic survey efforts of Severe Acute Respiratory Syndrome Coronavirus 2 (SARS-CoV-2) led to the emergence of over 200 distinct lineages using the Phylogenetic Assignment of Named Global Outbreak (PANGO) nomenclature system^2^. This includes the variants of concern (VOCs), such as Alpha, Beta, Gamma, Delta, and Omicron, that have been extensively studied^3–6^. Despite considerable research on viral evolution, the complex dynamics of viral competition during transmission across scales remain poorly understood.

Phylogenetic analysis has identified two major lineages of SARS-CoV-2 during the first three months of the pandemic outbreak, known as lineages A and B (or S and L)^2,7,8^. The lineages A and B are represented by genomes IME-WH01 and Wuhan-Hu-1, collected on 30 December 2019 and 26 December 2019, respectively^9,10^. Lineage A is considered ancestral to lineage B due to its closer genetic similarity to animal coronaviruses (Supplementary Fig. 1a)^2,7,11^, differing from lineage B by two single nucleotide variations (SNVs), C8782T (ORF1ab, C8517T, synonymous mutation) and T28144C (ORF8, T251C, nonsynonymous mutation L84S). Historically, lineage B was initially found in cases associated with the Huanan Seafood Market in Wuhan, China, and has since become the predominant lineage globally^2^. How and why lineage A was largely replaced by B (Supplementary Fig. 1b, c) in early 2020 is unknown^11,12^. The biological effects of these mutations remain poorly understood due to logistical and experimental challenges, hindering the explanation of their evolution^13–16^. Previous research has suggested separate introductions for the two lineages^17^, with lineage B being introduced earlier^18^. However, the potential adaptive mutations and the complex interactions between lineages A and B present challenges. Investigating the evolutionary and transmission dynamics of the early A and B lineages is essential for understanding the initial stages of the COVID-19 pandemic and for gaining broader insights into the early evolution and transmission patterns of future emerging pathogens, commonly referred to as Disease X^19,20^.

Mathematical modeling offers a useful tool to understand, identify, and predict the evolutionary and transmission dynamics of infective pathogens that have posed great challenges to public health worldwide^21,22^. Confronted with the rapid global spread of COVID-19, population models were most commonly developed, with the incorporation of new parameters and compartments for complex pandemic simulations^23–27^. However, many transmission models neglect viral competition and evolutionary dynamics^28,29^, failing to account for competitive interactions of multiple variants^30^. Human globalization also influences the spatial patterns of viral transmission^31^, with various social factors impacting the evolution and spread of viruses in human populations^32^. Furthermore, most models lack experimental validation, relying on theoretical derivations and simulations, limiting their predictive power and practical use^33^. To address these challenges, we aim to develop a succinct yet expandable infectious disease model to simulate real-world evolutionary epidemiology dynamics.

We tested the following theories regarding the driving forces of early evolutionary dynamics of COVID-19: 1. Host adaptation^34^: new mutations arise as pathogens adapt to human hosts, as seen with the influenza virus^35,36^. 2. Intra-host competition: co-infecting pathogens compete, influencing viral fitness and evolution^37^. 3. Inter-host competition: viral strains compete to infect the next host^38,39^. 4. Random genetic drift: most mutations are neutral, and random factors may drive the majority of evolutionary events^40^. Some studies suggested that lineage B might have been introduced to humans earlier than lineage A, leading to the founder’s advantages^18^.

In this study, we integrated global genomic surveys, early clinical samples, cell experiments, and mathematical modeling to analyze the evolutionary dynamics of SARS-CoV-2 from micro-to macro-scale. This multi-dimensional approach allowed us to investigate viral behavior from multiple perspectives, including adaptive mutations, intra-host competition, and inter-host transmissibility. Building on these insights, we developed the evoSIR-Seeding-Spreading (evoSSS) evolutionary infectious disease model, an innovative framework that incorporates both intra- and inter-host viral dynamics within a spatial epidemiological context. By integrating diverse data sources and offering a novel model to explore viral competition, our study provides a comprehensive understanding of how SARS-CoV-2 lineages competed and evolved. Beyond shedding light on the early dynamics of SARS-CoV-2, this work also provides valuable insights that can inform the study of emerging viral pathogens and enhance pandemic preparedness.

## Results

### Spontaneous mutations from lineage A to B were unlikely

We were curious why lineage A was rapidly overtaken by lineage B in early 2020. Despite lineage A’s closer genetic similarity to animal coronaviruses, strains of lineage B quickly became dominant after the outbreak, evolving into variants of concerns (VOCs). One possibility is that lineage B arose from lineage A through accumulating adaptive mutations in humans^34^.

To test this experimentally, we chose early lineage A and B strains sampled from six early COVID-19 patients for prolonged culturing in human cell lines to simulate the adaptive evolutionary process (Supplementary Fig. 2a, b)^41^. Three lineage A strains and two lineage B strains underwent three passages (72 hours each) in Calu-3 and Vero cell lines. Mutation spectra in the progeny of both lineages were aligned with previously reported results (Fig. 1a)^41^, and no significant nucleotide substitutions at loci 8782 and 28144 after three passages were found (Supplementary Fig. 2c). This result, showing no spontaneous mutations from lineage A to B, supports the absence of intermediate haplotypes (with one SNP instead of two) in early 2020 sequences^18^. Therefore, the evidence supported the hypothesis of multiple, independent zoonotic transmissions for the two lineages and laid the foundation for our subsequent analysis of the competitive infections and transmissions between lineages A and B.

**Fig. 1.**
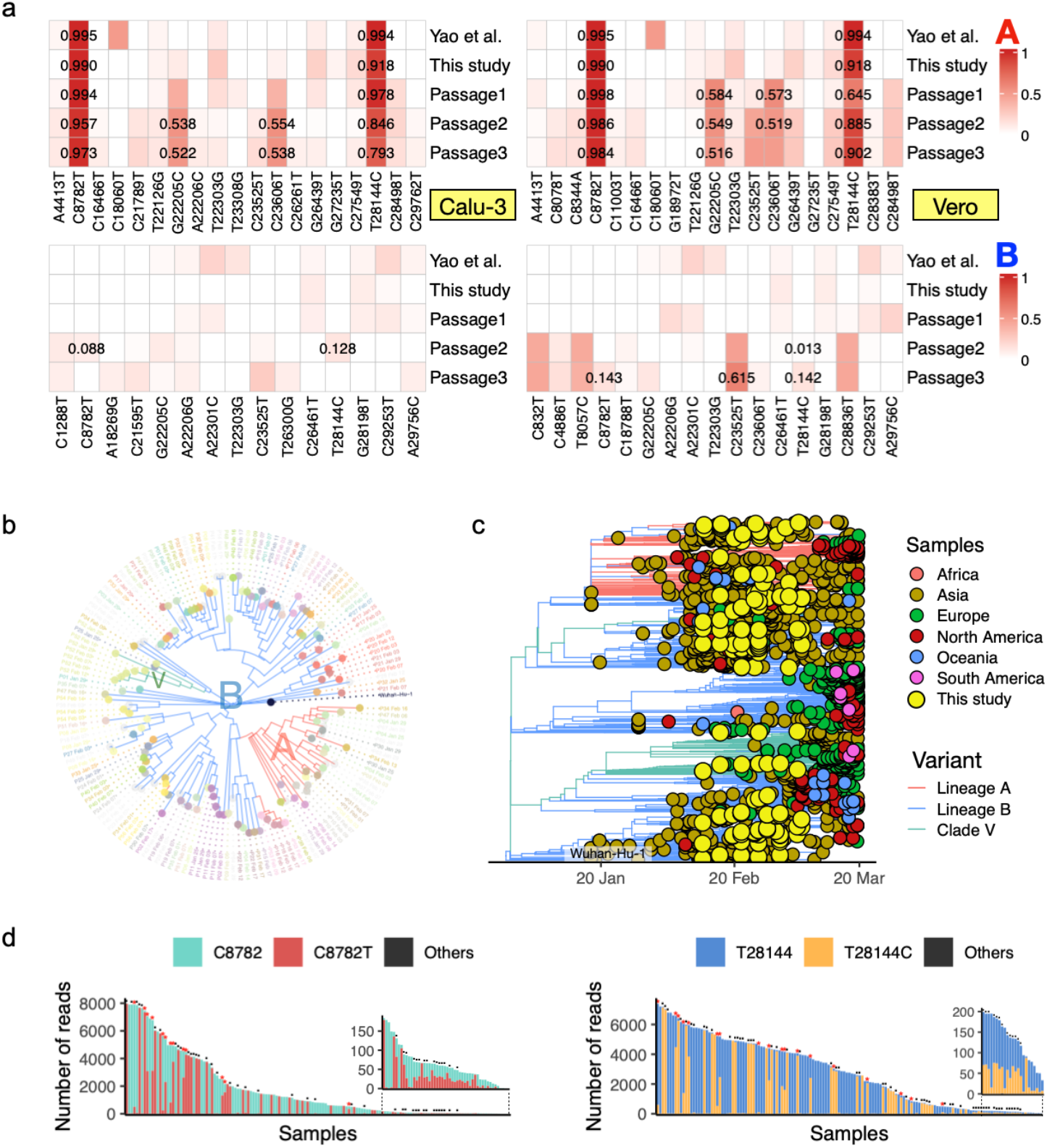
Co-infection of lineages A and B in early patients of COVID-19. **(a)** Consistent mutation map of lineages A and B across three 72-hour passages in Calu-3 and Vero cell lines. The top row displays infection experiments with lineage A strains, while the bottom shows those with lineage B strains. “Yao et al.” refers to data from a previous study^41^, and “This study” represents the initial sequencing data before the first passage in this study. Variant allele frequency (VAF) at loci 8782 and 28144, as well as variants with VAFs exceeding 0.5, are labeled. **(b)** Maximum likelihood phylogenetic tree of clinical viral genomes. Branches are colored according to lineages (A, B, and V), with each tip corresponding to an individual viral genome labeled with sample identifiers and collection dates. Patients with longitudinal samples are distinguished by unique colors, while samples with only a single timepoint are shown in light grey. **(c)** Maximum likelihood phylogenetic tree of early viral populations, constructed using data from this study and 2,081 sequences from the GISAID database as of March 1, 2020. Tips were colored by regions, and samples from the current study were highlighted in yellow. In (**b**) and (**c**), the central node marked with “Wuhan-Hu-1” (lineage B) represents the reference genome. **(d)** Stacked histogram of nucleotide distributions at sites 8782 and 28144, as determined by the number of mapped sequencing reads. Each bar represents a clinical sample in (**b**), with the height corresponding to the total number of sequencing reads for that sample. The colors in the bars indicate the proportions of different nucleotides at these sites. The black portions at the bottom represent nucleotides other than C or T, though these are minimal. Asterisks (*) in red signified a mixture of lineages A and B where both original and mutated nucleotides have at least 100 supporting reads, while dots (.) indicated a similar mixture but with at least 20 supporting reads. The varying heights of the bars represent differences in sequencing depth across samples.

### Prevalent co-infection of lineages A and B in early patients

To investigate the evolution and transmission of lineages A and B during the early COVID-19 outbreak, we collected 149 longitudinal samples from 66 COVID-19 patients admitted to Zhejiang University-affiliated hospitals between January 25, 2020, and February 17, 2020 (Fig. 1b). The viral sequences of our samples represent the earliest cases in SARS-CoV-2 evolution, encompassing lineage A and B strains, which were broadly distributed in sequence records from the GISAID database prior to March 1st, with most linked to Asia (Fig. 1c). In addition, the G26144T (G251V in ORF3a; observed in 21 patients) and G11083T (L3606F in ORF1a; observed in 9 patients) mutations were identified as marker variants of the ‘V’ clade, evolving from the early lineage B (GISAID). The Gamma lineage, recognized as a VOC nearly ten months later, exhibited the C13860T mutation (synonymous in ORF1b), observed in 21 patients (Supplementary Fig. 3a, b). The mutation spectrum of our samples indicated substantial genetic diversity during the initial outbreak period.

Interestingly, variant analysis of sequencing reads revealed significant nucleotide mixing at sites 8782 and 28144, suggesting many patients were co-infected with both lineages (Fig. 1d). This was likely due to superinfection (multiple sources of infection) and co-transmission^42,43^, as adaptive mutations were deemed unlikely based on both experimental (Fig. 1a) and epidemiological evidence^18^.

Notably, eight patients were also involved in a previous study on patient-derived mutation spectra during the initial outbreak^41^. The viral genomes from these patients showed consistent mutations across both studies, validating our data. However, a comparison of sputum and fecal samples from the same patients revealed significant differences in mutation spectra, likely due to distinct selective pressures in the gastrointestinal tract (Supplementary Fig. 3c)^44^.

### Intra-host competitive dynamics of lineages A and B

The prevalence of co-infection led to the hypothesis that the competitive advantage of lineage B *in vivo* may have contributed to its dominance. To test this, we designed competition assays commonly employed to assess the fitness of SARS-CoV-2 strains, such as Alpha versus Beta and D614 versus G614^45–47^. An important caveat in interpreting the changes in mutation frequency due to competition is the absence of intra-host adaptive mutations when grown independently (Fig. 1a).

We tested six combinations of lineages A and B in competition assays using Calu-3 and Vero cells in triplicate. Equal plaque-forming units (PFUs) of strains from both lineages were mixed and inoculated, confirmed by a balanced SNV representation of lineages A and B in the viral populations immediately post-inoculation (Supplementary Fig. 4a, b). Culture medium samples were collected for sequencing at 0, 8, 24, 48, and 72 hours post-infection, as well as after two passages. Surprisingly, contrary to expectations, lineage A strains demonstrated noticeable but not overwhelming superiority over B, as evidenced by the higher proportions of lineage A strains in multiple competition trials (Fig. 2a and Supplementary Fig. 5a). The competition profile differed between Calu-3 and Vero cells, with lineage A presenting a stronger competence in Vero cells.

**Fig. 2.**
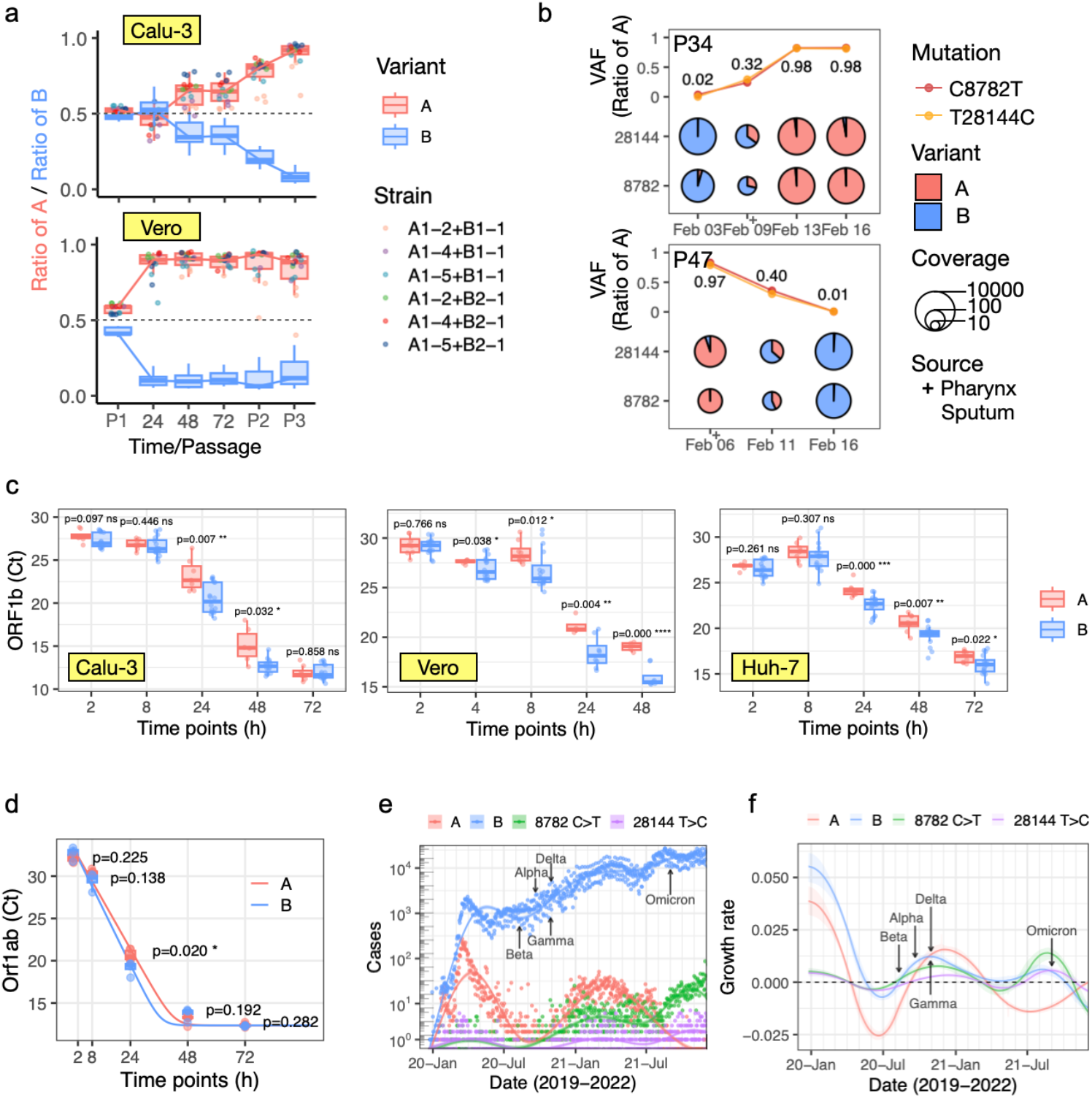
Competitive dynamics of lineages A and B in cell lines and early patients. **(a)** Competition experiments between lineages A and B in Calu-3 and Vero cells. Equal PFUs of lineage A and B strains were mixed and inoculated onto cell lines. The time-series plots depict the dynamics of lineage A and B ratios across two rounds of passages. Colored dots represent six different combinations of A and B strains, with each combination conducted in triplicate. **(b)** Longitudinal dynamics of lineages A and B within hosts P34 and P47. The charts display the dynamics of the lineage A ratio across longitudinal samples, represented by the VAFs of C8782T and T28144C. The size of the pie charts corresponds to the sequencing coverage, with larger pies indicating higher coverage. **(c)** Boxplots showing the replication dynamics of lineages A and B in different cell lines, based on data from the previous study^41^. The y-axis shows the cycle threshold (Ct) values of the SARS-CoV-2 ORF1b gene, with lower Ct values indicating higher viral titers. **(d)** Boxplots showing the replication dynamics of lineages A and B from infection experiments conducted in 2023. The replication dynamics were fitted using logistic growth regression. The p-values in (**c**) and (**d**) were calculated using the t-test at each time point. **p < 0.01, ***p < 0.001. **(e)** Temporal dynamics of lineage A, B, 8782 C>T, and 28144 T>C. The points represent the number of daily collected sequences in the GISAID database. The curves were estimated using a generalized additive model with penalized cubic regression spline (solid line), and 95% credible intervals were shown in shaded regions. The emergence of variants of concern (VOCs) is annotated on the curves. **(f)** Temporal dynamics of the daily growth rate of lineage A, B, 8782 C>T, and 28144 T>C. The daily growth rates were estimated from 1000 times bootstrapping. The dotted line indicates where the growth rate equals zero.

We reckoned that understanding the true impact of these mutations solely based on cell lines or even animal models might be challenging^48^. We thus delved deeper into the longitudinal dynamics of lineages A and B in early COVID-19 patients. Nine patients showed a change in mutation frequency greater than |0.1| at both the 8782 and 28144 loci over time (Supplementary Fig. 5b, c). In contrast to the consistent dominance of A in cell lines, the co-infection dynamics in patients were more variable. Longitudinal samples from the same patient showed instances where lineage A overtook B and vice versa, as observed in patients P34 and P47 (Fig. 2b). Overall, neither cell experiments nor clinical samples demonstrated a clear advantage for lineage B over A in intra-host competition.

### The faster replication rate of lineage B facilitated its transmission

To further explore the underlying advantageous traits of lineage B, we compared the replication dynamics of lineages A and B. Previously, we conducted cell infection experiments of early SARS-CoV-2 strains in Calu-3, Vero, and Huh-7 cell lines^41^, including early A and B strains (Supplementary Fig. 2a). We observed that strains of lineage B replicated significantly faster than lineage A at 24 and 48 hours post-infection (Fig. 2c), consistent with their higher infection ratios (Supplementary Fig. 5d). In 2023, three years later, we repeated the experiments using Calu-3 cells and found a consistent result, with lineage B replicating faster (Fig. 2d).

The epidemiological analysis revealed faster transmission for lineage B compared to lineage A during the initial outbreak phase (Fig. 2e), as evidenced by higher daily growth rate estimations derived from bootstrapping global sequenced cases (Fig. 2f and Supplementary Fig. 6a, b). The estimated exponential daily growth rates for lineages A and B were 0.039 and 0.055 (95% CI: 0.033-0.046 and 0.048-0.062), respectively, indicating a significant difference in their transmissibility.

Despite potential intra-host disadvantages in direct competition, the faster replication rate of lineage B might enhance its rapid transmission. The driving forces of evolution appeared to be more complex than replication or competition alone. These findings inspired the development of mathematical models to capture the intricate interplay of factors shaping viral evolution and transmission.

### Development of the evoSIR model for simulating competition and transmission dynamics of multiple variants

Emerging variants present a significant threat to public health, highlighting the need for robust modeling to monitor their spread^23,25,50–52^. While previous competitive models have described species interactions and viral competition during epidemics, they require refinement and integration to handle empirical data and large-scale pandemic scenarios^53,54^. To address this gap, we developed an evidence-based model that captures the complexities of competitive evolution and transmission dynamics in a highly mobile human society.

To simulate and understand the competitive interactions among variants, we developed the evolutionary-SIR (evoSIR) model, incorporating both intra- and inter-host competition components (Fig. 3a). The model was initially demonstrated with two competing variants, but it can be extended to multiple variants (Supplementary Fig. 7a). The intra-host component is based on the competitive Lotka– Volterra equation^55,56^, modeling the dynamic competition between two viral variants, V_1_ and V_2_, within a host. Key parameters include the mutation rate (μ), which describes the mutation process from V_1_ to V_2_, the replication rates of V_1_ and V_2_ (r_1_ and r_2_), and the inhibitory effects of V_1_ on V_2_ (a_21_) and V_2_ on V_1_ (a_12_) within the carrying capacity (K). Intra-host competition occurs when resources are limited.

**Fig. 3.**
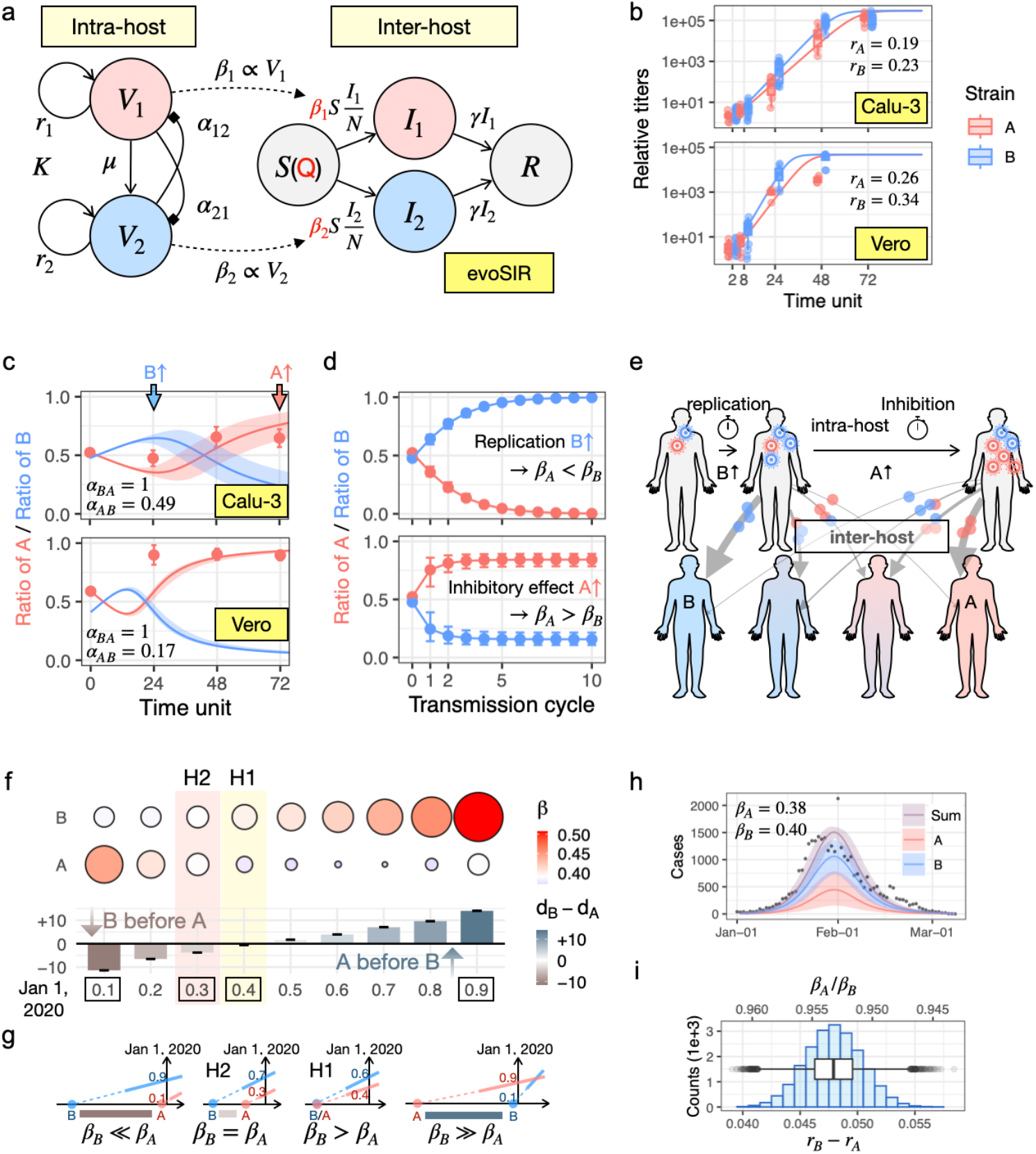
The evoSSS model depicted competitive dynamics intra- and inter-host. **(a)** Illustration of the evoSIR model. The evoSIR model incorporates the intra- and inter-host components of two competing variants, 1 and 2. In the intra-host component, V_1_ and V_2_ are the viral titers, and r_1_ and r_2_ are the replication rates of variants 1 and 2, respectively. K is the host’s carrying capacity for viruses. The a_12_ and a_21_ represent the inhibitory factors of variant 1 on variant 2 and variant 2 on variant 1. μ represents the evolution rate of viral mutation from V_1_ to V_2_. The intra-host component includes compartments for individuals infected with each variant. S is the number of susceptible individuals in proportion to the spreading capacity Q. I_1_ and I_2_ are the numbers of individuals infected by the two variants, and R is the number of removed individuals. The transmission rates β_1_ and β_2_ are positively correlated with the intra-host viral titers V_1_ and V_2_, and the removed rate *γ* was assumed to be the same for both variants. **(b)** The evoSIR model captures the titer dynamics of strains A and B in Calu-3 and Vero cell lines. The boxplots display the experimental data, calculated from Ct values as differences relative to the lowest titers in the samples. The lines represent the model simulations estimated using maximum likelihood estimation (MLE). **(c)** Fitting inhibitory factors (a_AB_) from the competition experiment (Fig. 2a). The inhibition from A to B (a_BA_) was fixed at one. Estimations were performed using Markov Chain Monte Carlo (MCMC). The two variants compete intra-host, with B initially replicating faster, while A exhibits stronger inhibitory strength, ultimately dominating. **(d)** Ratio dynamics of lineage A, starting at 0.5, over successive inter-host transmission cycles. The upper and lower panels depict two scenarios corresponding to the early (blue arrow) and later stages (red arrow) in (**c**). **(e)** Diagram depicting infection and transmission dynamics from intra-host to inter-host. Grey arrows represent inter-host transmission at the early and late stages of infection, with wider arrows indicating a higher likelihood of transmission for the dominant variants. **(f)** Relationship between relative introduction time and transmission rates of lineages A and B. The x-axis shows initial frequencies of lineage A (0.1 to 0.9) on January 1, 2020. The upper panel represents transmission rates (β), while the lower panel illustrates the relative introduction time difference (d_B_-d_A_), with negative values indicating lineage B appeared first. Shaded areas highlight hypotheses of faster transmissibility (H1) or earlier introduction (H2) for lineage B’s dominance. **(g)** Schematic representations of the growth dynamics of lineages A and B corresponding to (**f**) (black boxes). **(h)** The evoSIR model simulates the COVID-19 outbreak in Wuhan. The parameters of transmission and removal rates were estimated using MCMC to fit Wuhan data^23,49^. The estimated number of total infectious cases (Sum) and those infected by A and B were depicted. The colored points denote the mean values, and shaded areas represent 95% credible intervals. **(i)** Estimated distribution of the difference in replication rates (r_B_ - r_A_, the lower axis) corresponding to the estimated ratio of transmission rates (β_A_/β_B_, the upper axis) derived from (**h**).

The inter-host component was modeled as a competitive exclusion equilibrium, extending the classical SIR model^57^ to account for different variant infections. It includes susceptible individuals (S), whose proportion is influenced by the spreading capacity (Q), infectious individuals (I_1_ and I_2_) who are infected by V_1_ and V_2_, and removed individuals (R). Variants infected the same susceptible population, with the more competitive variant potentially driving others to extinction. The transmission rates (β_1_ and β_2_) were assumed to be positively correlated to the proportions of each variant’s titer (β_i_∝V_i_/(V_1_+V_2_)). We assumed identical removal rates *γ* for both variants, as there are no observed differences in clinical outcomes between them^58^.

The parameter values for the intra-host component were estimated using maximum likelihood estimation (MLE)^59^ from infection experiments in Calu-3 and Vero cell lines (Fig. 2a). The inter-host component parameters were estimated using Markov Chain Monte Carlo (MCMC) fitting of Wuhan daily incidences^23,49,60^.

### The evoSIR model delineated the competitive evolution of lineages A and B intra- and inter-host

The evoSIR model integrates intra- and inter-host components to bridge experimental findings with epidemiological dynamics, simulating the progression from host infection to viral transmission. We set the mutation rate to 1e^-6^ per base per replication cycle based on the estimated mutation rates per replication cycle for SARS-CoV-2 and other betacoronaviruses^48^. In the single-variant simulation, the variant remained stable, consistent with infection experiments (Supplementary Fig. 7b). While lineage B exhibited a faster replication rate (r_B_>r_A_) in infection experiments (Fig. 3b), the fittings of competition experiments showed a stronger inhibitory effect by lineage A (a_BA_>a_AB_) (Fig. 3c and Supplementary Fig. 7c). Despite lineage B’s higher replication rate, lineage A dominated due to its stronger inhibitory effects. In some instances, the dominance of one lineage depended on the balance between faster replication or stronger inhibition (Fig. 3d, e). The proportion of variants with competitive traits increased through successive transmissions, driving higher transmission rates in epidemic hotspots. Real-world data showed that transmission often preceded symptoms^61^, highlighting the role of early viral dynamics over inhibitory effects in resource-limited environments.

To explore alternative evolutionary scenarios underlying the quick dominance of lineage B, we performed a sensitivity analysis using Wuhan data^23,49^, varying the initial proportion of lineage A infections (p_A_ from 0.1 to 0.9) on January 1, 2020, aiming for 30% prevalence of lineage A (Fig. 3f, g and Supplementary Fig. 8a, b). In the first hypothesis (H1), both variants were introduced simultaneously, with lineage B exhibiting a faster transmission rate (Fig. 3h). Assuming exponential correlation between transmission and replication rates, replication rate differences (0.042-0.055) from inter-host modeling aligned with those from intra-host modeling (Fig. 3b, I). The second hypothesis (H2) proposed that lineage A was introduced after lineage B, with identical transmission rates^18^. This hypothesis estimated a two-day delay for lineage A but did not fully explain its rapid decline, considering its early global prevalence in regions such as China, Spain^62^, the US^63^, Thailand, and the United Arab Emirates before March 1, 2020 (Supplementary Fig. 9a, b). Therefore, deterministic factors, such as replication and infection capabilities, were critical in driving the competitive transmission dynamics between viral lineages, particularly in the case of lineage B’s dominance over lineage A.

### The evoSSS model simulated competitive transmissions of a large-scale pandemic in a mobile human society

To extend modeling from local epidemics to a global pandemic, we integrated the evoSIR model with the Seeding-Spreading framework, creating the evoSIR-Seeding-Spreading (evoSSS) model. This framework treats the pandemic as a series of local propagation cycles (Fig. 4a). We used superscripts to denote the cycle number, starting with the initial outbreak (I^0^) at cycle 0. Infected travelers (D^n^) from each wave of spreading (I^n-1^) act as seeding populations, initiating new hotspots (h_1_, h_2_, …, h_j_) and interacting with susceptible individuals to form the next wave of infections (I^n^). The seeding populations (D^n^) are generated by the seeding function f_seed_(M) (where M is the mobility matrix), spreading diseases in new hotspots via the evoSIR model (f_evoSIR_(Q), where Q is the spreading capacity generating the susceptible population) (Fig. 4b). Incorporating mobility (M) and spreading capacity (Q) allows the model to account for social interventions such as travel restrictions, government policies, and vaccination campaigns. The outcomes from all hotspots aggregate to represent the global pandemic state I^n-1^→I^n^.

**Fig. 4.**
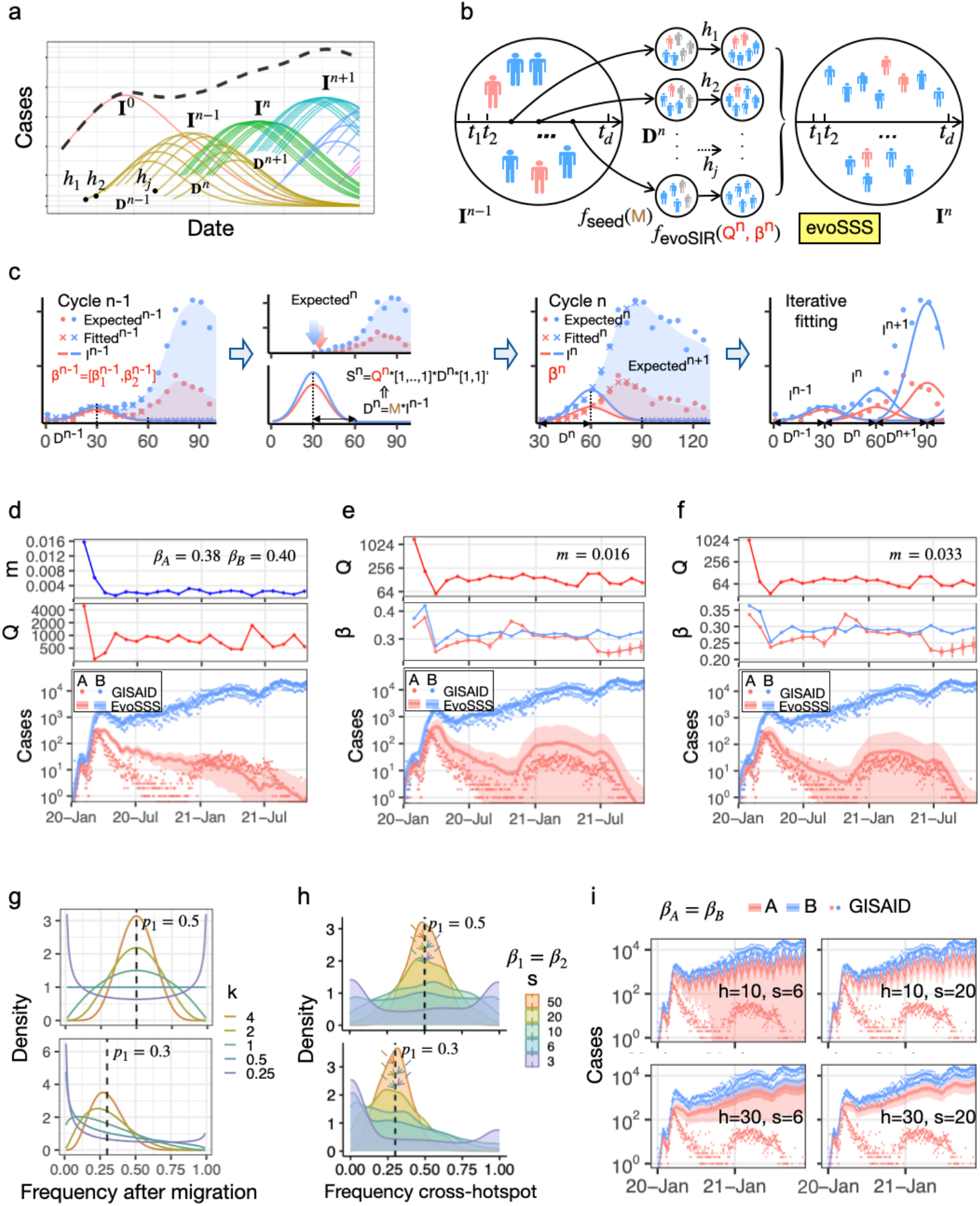
The evoSSS model simulated large-scale pandemics of prevailing variants in mobile human populations. **(a)** Illustration of the Seeding-Spreading transmission framework. Different colors of curves indicate epidemic states of different cycles. The black dashed line illustrates the observable total number of infections, showing the transition from localized outbreaks to a global pandemic. **(b)** Illustration of the evoSSS model. The evoSIR-Seeding-Spreading (evoSSS) model integrates the evoSIR model with the Seeding-Spreading framework. **(c)** Illustration of the iterative fitting process. The seeding population, derived from the susceptible population of the previous cycle, triggered outbreaks in new hotspots, driving the propagation of the epidemic state iteratively from cycle n-1 to n. **(d)** Simulations of large-scale COVID-19 pandemics using the evoSSS model with constant transmission rates (β_A_=0.38, β_B_=0.40) fitted from Wuhan data (Fig. 3e). The dots represent GISAID counts. The lines denote the mean values, and shaded areas represent 95% credible intervals of the evoSSS simulations. The top two panels illustrate the dynamics of the estimated mobility values and transmission rates for each cycle. **(e and f)** Simulations of large-scale COVID-19 pandemics using dynamic transmission rates for lineages A and B. The mobility values were fixed at 0.016 (**e**) and 0.033 (**f**) across all cycles. **(g)** Theoretical distribution of variant frequencies among population subdivisions with initial ratios p_1_ = 0.5 and p_1_ = 0.3. The theoretical distribution of frequency p after migration follows: y(p) = C * q^4kp1-1^ * (1-q)^4k(1-p1)-1^, where k (bottleneck coefficient) correlates to the seed population^64^. **(h)** Simulated distribution of frequencies of V_1_ (x-axis) during across-hotspot transmission with small seed populations (s). The mean values (dashed lines) in the simulations were approximately the initial frequencies (p_1_ = 0.5 and p_1_ = 0.3). **(i)** Bottleneck effects in the global transmission of lineages A and B were limited. The wide shades represent the 95% credible interval, while the narrow ribbons show the 50% credible interval. The four panels illustrate how bottleneck effects are influenced by the number of hotspots (h) and seed populations (s).

The evoSSS model used MCMC for iterative fitting, with priors derived from the previous cycle (Fig. 4c). Specifically, we fixed 30 hotspots and 30-day seeding-spreading cycles. The mobility matrix (M) was diagonal with a default 30-length vector (m, …, m)_30_ of mobility value (m), and the removal rate (*γ*) was 0.157 based on Wuhan data. The spreading capacity (Q) and transmission rates (β=[β_1_,β_2_]) for the two variants were estimated using inter-host evoSIR modeling over 100 days, with a zero constraint applied after two cycles (60 days). For subsequent cycles, new infectious individuals were calculated by subtracting the simulated onset cases from the previous cycle. Infectious seeds generated from the previous cycle (D^n^=M*I^n-1^) triggered outbreaks in new hotspots, where the size of the susceptible population was determined by the spreading capacity (S^n^=Q^n^*[1,..,1]_30_*D^n^*[1,1]). The model fitting performance was assessed by checking the effective sample size (ESS>100) and the potential scale reduction statistic 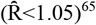.

We applied the evoSSS model to large-scale pandemic data for lineages A and B. The initial state was derived from Wuhan cases with fixed transmission rates (β_A_=0.38, β_B_=0.40) (Fig. 3h), fitting the parameters capacity (Q) and mobility (m) after scaling GISAID data for Wuhan cases. The simulation accurately reflected the global COVID-19 pandemic, showing a continuous decline in lineage A and quick dominance of lineage B (Fig. 4d). Dynamic fitting of transmission rates, with fixed mobility values to prevent overfitting, captured the resurgence of lineage A before its eventual disappearance (Fig. 4e, f). Faster transmission rates of lineage B led to its quick dominance in the global pandemic. Furthermore, the mild increase in lineage A under low mobility conditions suggests that the stochastic effect can counteract its intrinsic fitness advantage (Fig. 4e).

### Stochastic bottleneck events were mitigated in a highly mobile society

Both deterministic and stochastic factors are critical for understanding pathogen phylodynamics^66^. Stochastic factors, such as bottleneck events, occur when a small number of infected individuals seed a new hotspot. These events were observed during the early transmission stages of both lineages A and B, exhibiting distinct regional distribution patterns (Supplementary Fig. 9a, b). This phenomenon can be described mathematically by the distribution equation y = C * q^(4kp1-1)^ * (1-q)^4k(1-p1)-1^, where p_1_ is the variant frequency in the original population, and bottleneck coefficient k correlates with the size of the founding population (Fig. 4g)^64^. When k < 0.5, one variant tends to fix over others.

The evoSSS model effectively simulated bottleneck effects during the transmission of two variants to new locations, starting with initial proportions of 0.5 and 0.3 for variant 1 (Fig. 4h). Spatial bottlenecks were captured with small seed populations in new hotspots during disease spreading. A tendency for one variant to dominate was observed with seed populations of 3-6, resembling typical family sizes. This suggests bottlenecks may occur when infected families travel. However, these local stochastic effects are reduced in a highly mobile global society, where larger traveling seed populations and numerous epidemic hotspots reduce bottleneck influence (Fig. 4i and Suppplementary Fig. 10a).

During the early stages of the COVID-19 pandemic, the local and global evolution of lineages A and B was influenced by the interplay between transmission rates and bottleneck events. Transmission rates, acting as deterministic traits, consistently favored lineage B by reducing the proportion of lineage A. In contrast, bottleneck events introduced stochastic variation, leading to significant fluctuation in local proportions of each variant. Despite local stochastic effects, the intrinsic advantages of specific variants remained the primary driver of global viral evolution.

### The evoSSS model predicted competitive transmission dynamics of various infectious diseases

To address potential future epidemic outbreaks, we aim to use the evoSSS model to predict the evolution and transmission dynamics of infectious diseases under various scenarios. Spreading capacity, influenced by factors such as social interventions, seasonal outbreaks, and the nature of the virus, limits the number of susceptible individuals (Fig. 5a). Transmission rates, representing the competitive strength of the variants, serve as early warnings for potentially high-risk variants (Fig. 5b).

**Fig. 5.**
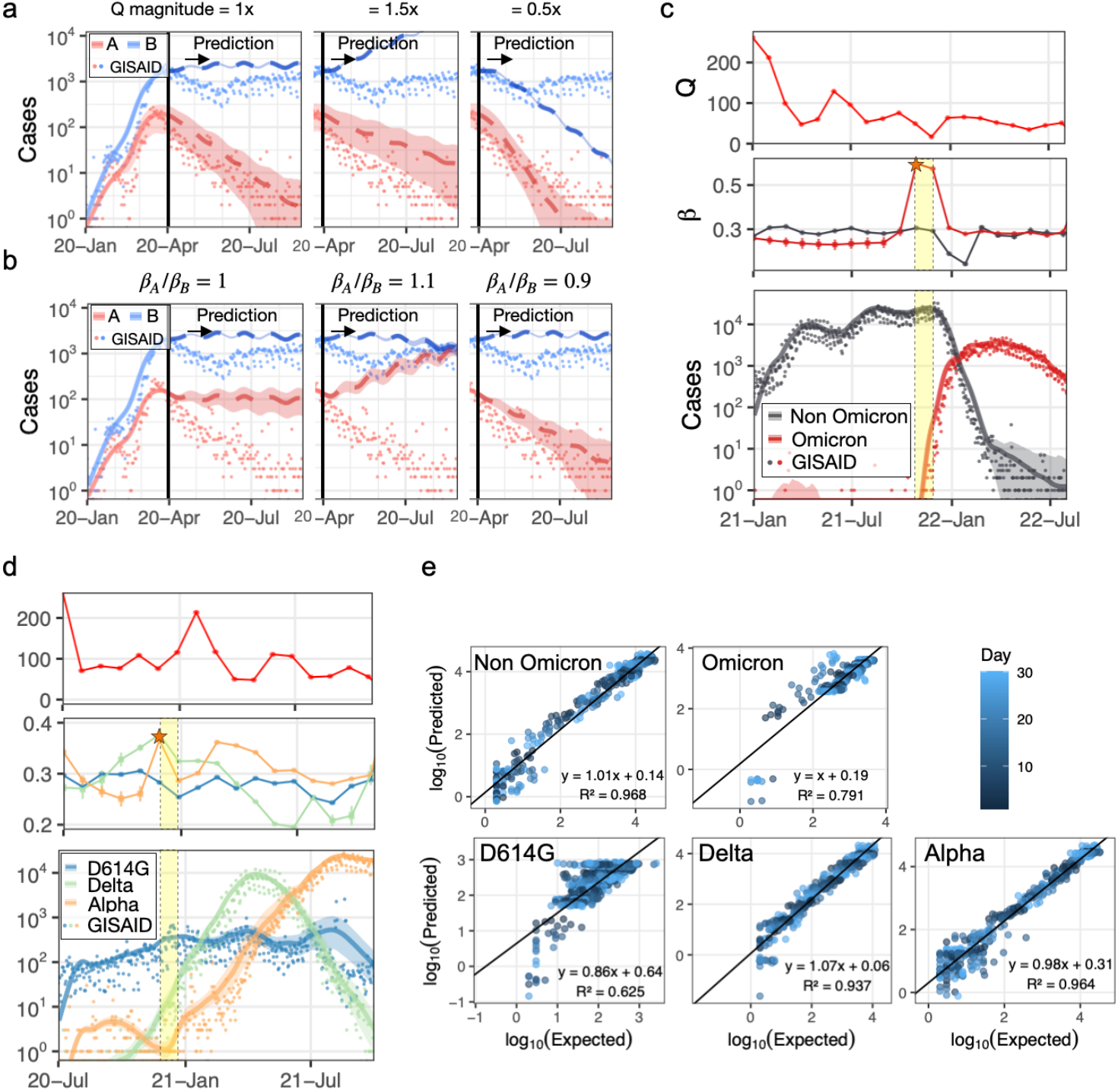
The evoSSS model predicted competitive transmission dynamics of SARS-CoV-2 variants. **(a-b)** Predictions of the competitive dynamics of A and B under varying spreading capacity (**a**) and transmission rate ratios (**b**). The other parameters are set as in the final transmission cycle fitting. The vertical black line marks the prediction start point, with shaded areas indicating 95% credible intervals. **(c-d)** Simulations of Omicron vs. non-Omicron variants (**c**) and D614G, Alpha, and Delta variants (**d**) during COVID-19. Spreading capacity (Q) and transmission rates (β) are shown, with stars marking high-risk transmission points of potential variants and yellow areas highlighting peak transmission periods. **(e)** Predictions of SARS-CoV-2 variants from (**c**) and (**d**). Each dot represents a prediction for a specific number of days ahead, indicated by the color gradient. The x-axis shows the logarithm of recorded cases, and the y-axis shows the logarithm of predicted cases. Linear fitting equations and R^2^ values are annotated.

We first applied the evoSSS model to simulate the competition between Omicron and non-Omicron variants (Fig. 5c) and between D614G, Alpha, and Delta variants (Fig. 5d). For Omicron, transmission rates increased notably when daily cases were below a hundred, serving as an early alert for a potential outbreak. Similarly, early transmission rate peaks in Alpha and Delta variants signaled the emergence of variants of concern (VOCs). Using spreading capacity and transmission rates from the previous cycle, the model provided one-month-ahead predictions, demonstrating its capability for real-time surveillance of prevailing variants (Fig. 5e).

Importantly, the evoSSS model naturally extends beyond SARS-CoV-2 research. We applied it to model influenza and monkeypox epidemics, as well as the interaction between influenza virus and SARS-CoV-2, using monthly case counts from WHO FluNet^67^ and the GISAID database. Transmission rates and spreading capacities for influenza A and B were estimated by fitting the monthly total cases with daily averages of observed and simulated counts (Fig. 6a). The model effectively captured seasonal influenza outbreaks, with spreading capacity peaking annually. Using periodic spreading capacities, it predicted weekly and monthly dynamics, as well as the transmission patterns of influenza A and B for the following year (Suppplementary Fig. 10b). The model also simulated the transmission dynamics of influenza subtypes in recent years, highlighting the growth of transmission rates for A.H5 and the potential for its dominance in future seasons (Fig. 6b). For the monkeypox pandemic, we simulated the transmission dynamics of clades I and II using a limited dataset from the GISAID database. The results highlighted the risk of clade I in the monkeypox virus and emphasized the need for stronger surveillance and data collection (Fig. 6c). Overall, the simulations aligned with the reports on the increasing diversity and pathogenicity of the bird flu virus A.H5^68,69^ and monkeypox clade I^70,71^, demonstrating the evoSSS model’s potential in managing disease X in public health.

**Fig. 6.**
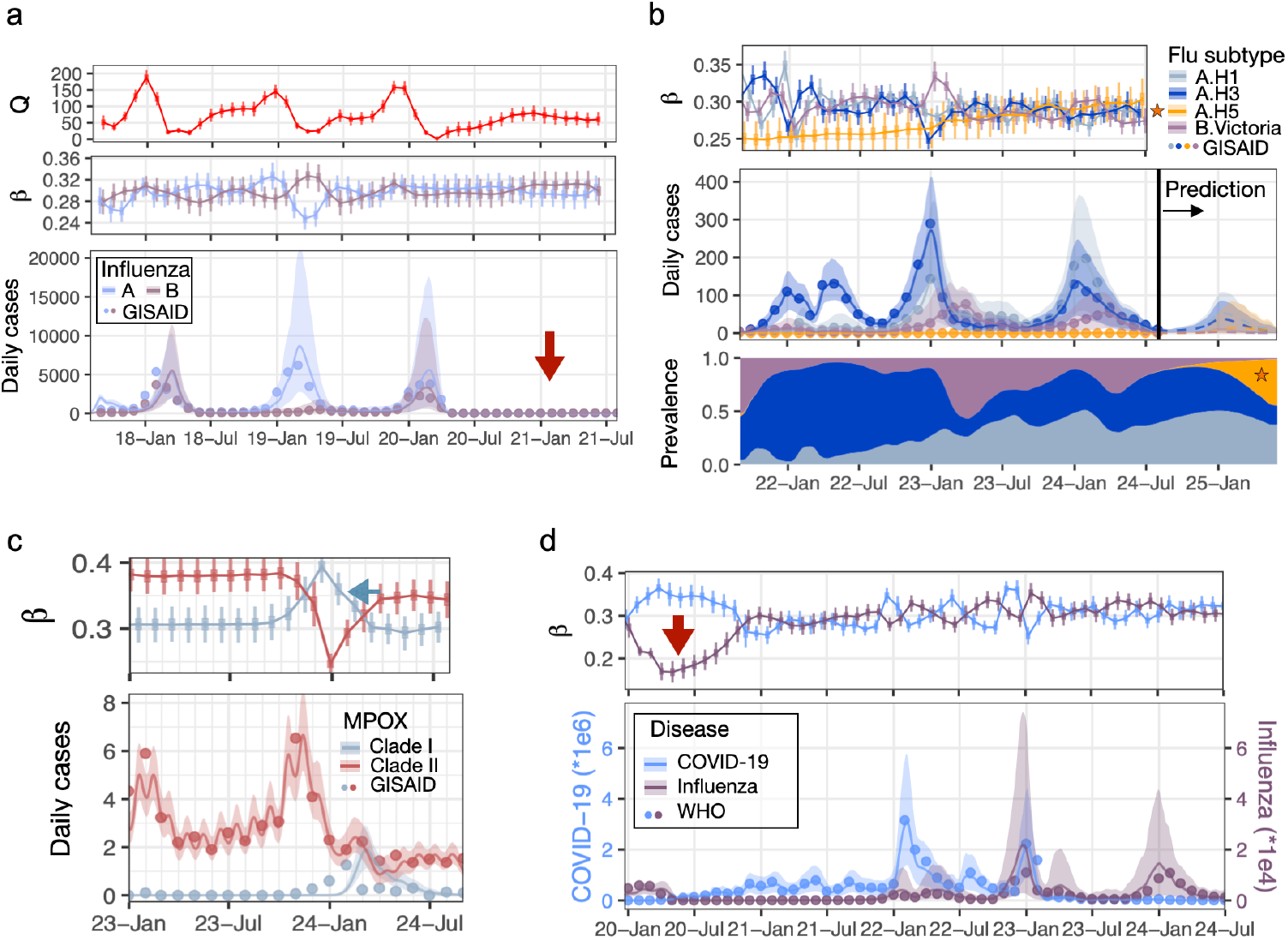
The evoSSS model captured variant risks and interactions in infectious disease dynamics. **(a)** Simulations of influenza A and B dynamics. Dots represent daily average cases calculated from monthly WHO FluNet data^67^. The evoSSS model parameters were fitted to match the monthly cases. **(b)** Simulations of the growth dynamics of influenza subtypes, with predictions in dashed lines starting from August 2024. The bottom panel shows the simulated and predicted prevalence of influenza subtypes, highlighting the potential prevalence of A.H5. **(c)** Simulations of monkeypox. The green arrow highlights the upward trend in the transmission rates of clade I. **(d)** Competitive dynamics between influenza and COVID-19. The red arrows in (**a**) and (**d**) indicate the supposed inhibitory period of influenza.

While modeling and predicting influenza dynamics, we observed an unusual inhibitory period starting in 2020, coinciding with the onset of the COVID-19 outbreak (Fig. 6a and Suppplementary Fig. 10c). The evoSSS model captured the significant suppression of influenza transmission rates alongside the rise of SARS-CoV-2 transmission during this period (Fig. 6d). The first notable influenza wave after the COVID-19 outbreak, from 2021 to 2022, exhibited a wide ‘M’-shaped transition, likely influenced by the COVID-19 outbreak (Suppplementary Fig. 10d). These unusual influenza patterns after 2020 were likely driven by its interaction with SARS-CoV-2 and social intervention and COVID-19-related social interventions^72–75^.

## Discussion

Our study focuses on infectious disease modeling and offers valuable insights into the competitive evolution of prevailing pathogens in the global spread of pandemics. Based on the clinical, experimental, and epidemiological evidence of SARS-CoV-2, we developed the evolutionary-SIR-Seeding-Spreading (evoSSS) model to enhance our understanding and guide more effective control strategies in the management of Disease X. The evoSSS model integrates the evoSIR model within the Seeding-Spreading framework, offering flexibility by allowing replacement of evoSIR with other spreading models. The evoSSS model offers a comprehensive framework to simulate and predict competitive dynamics of multiple pathogenic variants during a large-scale pandemic, bridging key aspects of disease modeling such as evolution, transmission, intra- and inter-host dynamics, and both deterministic and stochastic factors. Validation of the model using independent real-world cases from influenza and monkeypox pandemics confirms the evoSSS model as a robust, versatile tool for responding to emerging infectious threats.

To ensure the reliability of our research, we have conducted preparatory studies focusing on COVID-19 research, including optimization of bioinformatics analysis, experimental serial dilution tests across different sequencing approaches, and investigation of the phylodynamics of lineages A and B (Suppplementary Fig. 11-13 and Supplementary Text). Our analysis of clinical, experimental, and epidemiological data revealed complex interactions and competitive dynamics between SARS-CoV-2 variants. The conservation of mutations in lineage A and its competitive advantage challenge the assumptions that adaptive evolution or intra-host competition drives lineage B’s dominance. Instead, the difference between competition experiments and clinical data (Fig. 2a, b) suggests an influence of individual immunity. Similarly, in animal models, both Alpha and Delta variants outcompeted the fast-transmitting Omicron BA.1 variant^76^. Strain adaptation may confer benefits in replication and transmission while resulting in milder clinical outcomes. Among our samples, only two severe cases requiring ECMO were infected by lineage B, with no significant clinical differences between lineages A and B in the remaining cases. A study of 4,566 patients showed a milder outcome linked with lineage A despite its declining prevalence compared to B^77^. Another study with 326 patients in Shanghai suggested that host-dependent factors might outweigh genetic variation between lineages A and B^58^. Furthermore, the absence of adaptive evolution at the two sites in the early COVID-19 pandemic was consistent with the relatively slow evolutionary rate observed before the emergence of VOCs such as Alpha, Beta, and Gamma, which occurred approximately 8 months after the initial outbreak^48^. These observations supported the hypothesis of multiple spillover events^18^ or co-transmission of lineages A and B variants from animal hosts. Based on the results of infection experiments using early strains in multiple cell lines and the observed faster epidemiological growth rate of lineage B, we propose that its faster replication rate facilitates its rapid transmission.

Mixed infections were also observed in other studies, including Omicron and Delta variants^43^ and household co-infections^78^, providing the basis for the prevalent intra-host competition of variants. Faster intra-host replication and infection rates could significantly contribute to a more efficient spread of the advantageous variant over the other. Inspired by these intricate intra- and inter-host interactions, we developed the evoSSS model to simulate and better understand the competitive interactions among infectious pathogens. By fitting empirical data and parameter inspection, the evoSSS model effectively simulates the competitive dynamics of lineages A and B across scales, from intra-host to inter-host and from local outbreaks to global transmission. Furthermore, the model also offers a tool for exploring phenomena that are difficult to assess through empirical experiments^51^. Using this model, we explored the evolutionary mechanisms underlying competitive evolution. We show that faster replication in the early stages of infection facilitated rapid inter-host transmission; deterministic competitive factors drove global viral evolution in highly mobile human populations, while stochastic processes, such as bottleneck events, shaped local evolution.

We also demonstrate the capability of the evoSSS model as a powerful tool for predicting threatening variants and outbreak risks, offering valuable insights for public health management. Beyond coronavirus, viruses such as influenza, monkeypox, Zika, and Ebola require robust public health surveillance programs^51,52,79^. Moreover, the evoSSS model can effectively simulate the evolution and transmission dynamics of different viral pathogens concurrently. The marked suppression of the influenza virus transmission rates during the onset of the COVID-19 outbreak provided valuable insights into the interactions between influenza and SARS-CoV-2^72–75^. Moreover, the model should be applicable to infectious bacterial pathogens, given sufficient data.

Our study has some limitations, particularly in exploring the complexities of the COVID-19 outbreak. A key limitation is the need for high-quality longitudinal samples from the pandemic’s early stages. We did not investigate the molecular implications of mutations at sites 8782 and 28144 due to logistical and experimental challenges. Unlike well-studied mutations in the spike protein, which have been shown to improve receptor-binding capability^80^ and immune evasion^81^, the precise biological effects of these mutations remain unclear^13–16^. Additionally, the lack of sufficient epidemiological data for infections like monkeypox, along with delays and gaps in laboratory testing, poses challenges for further application of the evoSSS model. Mathematical modeling challenges, such as non-identifiability issues and complex real-world interactions, also need to be addressed^82^. In our study, the seeding process was represented using a simplified diagonal mobility matrix with a fixed number of hotspots across seeding-spreading cycles, allowing for manageable modeling of real-world complex scenarios. Furthermore, we did not use stochastic sampling simulations for each spreading cycle to reduce computational costs. Future advancements in modeling techniques and improved data quality will provide deeper insights into the evolution and transmission dynamics of infectious pathogens, as well as public health surveillance^30,51^.

By integrating empirical data with extensive computational modeling, our research endeavors to unravel the multifaceted evolution and transmission dynamics of the pandemic pathogens, from intra-host to inter-host and regional to global. This work provides valuable insights and a foundational framework for predicting and mitigating future outbreaks of Disease X.

## Methods

### Cell culture

The Vero (African green monkey kidney, ATCC® CCL-81™) and Calu-3 (human lung adenocarcinoma, ATCC® HTB-55™) cell lines were cultured in Dulbecco’s Modified Eagle’s Medium (DMEM; Gibco BRL, Grand Island, NY, USA) supplemented with 10% fetal bovine serum (FBS; Gibco BRL). The cells were maintained in a 5% CO_2_ incubator at 37°C. After treatment, cells were harvested and passaged using 0.25% trypsin and 0.02% EDTA.

Cell counts were performed using the Vi-CELL XR (Beckman Coulter, Miami, FL, USA), and cells were seeded at a density of 1 × 10^5^ cells/well in 24-well plates, one day (for Vero cells) or two days (for Calu-3 cells) prior to viral infection, to reach 80%–90% confluency.

### Human cohort

A total of 149 specimens were collected from 66 patients admitted to Zhejiang University-affiliated hospitals between January 25, 2020, and February 17, 2020. The specimens included 130 sputum samples, 10 pharyngeal swabs, five fecal samples, and four bronchoalveolar lavage fluid (BALF) samples.

The study was approved by the Clinical Research Ethics Committee of The First Affiliated Hospital, School of Medicine, Zhejiang University (Approval notice 2020-29) for emerging infectious diseases. All samples, including sputum, nasopharyngeal swabs, and stool, were collected with consent from COVID-19 patients. The samples were transported to a biosafety level III laboratory for viral isolation within 4 hours. The biosafety level was approved by the China National Accreditation Service for Conformity Assessment (CNAS No. BL0022), State Key Laboratory for Diagnosis and Treatment of Infectious Diseases, The First Affiliated Hospital, School of Medicine, Zhejiang University.

### GISAID data

The GISAID data (accessed on October 8, 2022) was retrieved from https://gisaid.org/, containing approximately 5.5 million SARS-CoV-2 sequences from human hosts, as well as environmental and animal sources. The data included whole genome sequences along with metadata, which met specific criteria such as completeness, high coverage, and a confirmed collection date. Coronaviruses originating from bats and pangolins were classified as SARSr-CoV for analytical purposes. Among the daily COVID-19 cases, the dataset included 11,646 complete genomes from lineage A, 5,422,021 from lineage B, 4,560 genomes exhibiting the 8782 C>T mutation, and 765 genomes identified with the 28144 T>C mutation.

### Wuhan data

The daily incidence of COVID-19 in Wuhan, spanning from December 8, 2019, to March 8, 2020, was obtained from previous studies^23,49^ and included 32,583 individuals who tested positive for SARS-CoV-2 based on laboratory-confirmed results.

### Additional data collection on influenza, monkeypox, and COVID-19

The data on influenza and monkeypox subtypes were obtained from GISAID, accessed on August 6, 2024, and September 4, 2024, respectively. The epidemiological data for influenza and COVID-19 over the entire period were sourced from the WHO, accessed on September 4, 2024.

### Viral infection and competition in cell lines

The first-round infection was conducted for both infection and competition assays. For infection assays, 100 TCID50 doses of viruses from three lineage A isolates and two lineage B isolates were inoculated onto 12-well plate cell cultures in duplicate. For competition assays, a mixed inoculation of equal doses of lineage A and lineage B viruses was used, resulting in six different combinations tested in triplicate. After a 2-hour incubation, cells were washed three times with PBS to remove non-binding viruses and replenished with 2 mL of fresh medium. At each time point (0, 8, 24, 48, and 72 hours) post-infection, 150 μL of culture supernatant was collected for RNA isolation using an automated magnetic bead-based method (MVR01, Liferiver Biotech, Shanghai), with 150 μL of fresh medium added to maintain the culture volume.

The following passages were conducted to test mutation consistency. For the second-round infection, two μL of the 72-hour culture supernatant from the first round was inoculated into fresh cell cultures. At 72 hours post-infection, 150 μL of supernatant was again collected for RNA extraction. The third-round infection followed a similar protocol, using the second-round supernatant at 72 hours post-infection for inoculation.

All RNA samples were stored at −80°C for parallel analysis. Upon thawing, the frozen samples from each timepoint were homogenized by pipetting and subjected to automated nucleic acid extraction (EX3600; Liferiver Biotech, Shanghai, China) using a magnetic bead-based kit (Cat No. MVR01; Liferiver Biotech). SARS-CoV-2 RNA quantification was performed on the CFX384 Real-Time PCR system (Bio-Rad) using the One-Step RT-PCR kit (ZC-HX-201-2, BioGerm, Shanghai), which simultaneously detects the viral ORF1ab and N genes. The viral samples from infection and competition assays at each time point were further processed for quantification and amplicon sequencing.

### RNA extraction, library construction, and sequencing

The total RNA in each deactivated clinical sample was extracted using a viral RNA mini kit (Qiagen, Germany). The extracted RNA was reverse transcribed using ProtoScript II Reverse Transcriptase (NEB, Cat. M0368), with 9 μL of the reaction product used for subsequent multiplex PCR experiments. Sequencing libraries were generated using the MultipSeq® SARS-CoV-2 Research Assay (iGeneTech, Beijing, China) according to the manufacturer’s instructions, and index codes were added to each sample. Qualified libraries were sequenced on Illumina NovaSeq 6000 with a pair-end 150 sequencing strategy.

### Statistical analyses and visualization

The majority of statistical analyses and visualization were performed using the Rstudio server (2021.9.0.351) and R (4.1.1). Additional support was provided by customized Python (3.8.13) scripts and Linux shell scripts. The primary R packages utilized were mostly from CRAN (https://cran.r-project.org/) and the Bioconductor project (https://www.bioconductor.org/). The essential packages used were ggplot2 (3.3.5), ggtree (3.2.1), ggtreeExtra (1.4.2), treeio (1.18.1), dplyr (1.0.7), tidyverse (1.3.1), RColorBrewer (1.1–2), scales (1.2.0), ggrepel (0.9.1), ggpubr (0.4.0), and stringr (1.4.0). In general, parametric statistical tests (t-test) were employed when the data was normally distributed (e.g., qRT-PCR measurements). The Bayesian inference and modeling were performed using rstan (2.21.8) and deSolve (1.35). The fitting was estimated using a generalized additive model with a penalized cubic regression spline, and the 95% credible intervals were represented as shades. The prevalence of strains was estimated using a binomial test, with 95% credible intervals displayed as error bars.

### Single-nucleotide polymorphism (SNP) analysis

Amplicon sequencing of viral genomic RNA was performed on the Illumina Novaseq 6000 platform, generating an average of 1.7 million post-cleaning reads per sample (average coverage exceeding 8,000×). The sequencing Fastq files were processed under quality control, filtering, and trimming of raw reads using fastp (0.20.1)^83^. The cleaned reads were then mapped against the SARS-CoV-2 reference genome Wuhan-Hu-1 (NC_045512.2) using bowtie2 (2.3.5.1)^84^. All mapped reads were extracted using samtools (1.9)^85^ with parameters “-F12 −q30”. Primer sequences were trimmed using BAMClipper (1.1.1)^86^ for amplicon sequencing data. Variants were called using VarScan2 (2.3.9)^87^ with parameters “--output-vcf 1 -min-reads2 10 --min-var-freq 0.05 --strand-filter 0”. The coverage was calculated using the mpileup file and the samtools mpileup command.

### ViroDecode

We developed the viral analysis workflow within the miniconda3:4.8.2 Docker environment (https://hub.docker.com/r/continuumio/miniconda3, accessed on January 10, 2023). ViroDecode was composed of two main functions: SNP calling and genome assembly. The workflow was optimized and standardized for ultra-deep sequencing and amplicon sequencing data, with a focus on research on the SARS-CoV-2 virus (Suppplementary Fig. 11). The filtered bam files from the SNP calling workflow were used for genome assembly. The alignment tools bwa-mem (0.7.17-r1188)^88^ and bowtie2 (2.3.5.1)^84^ were both available for use. For efficient genome assembly by SPAdes (3.13.0)^89^, a downsampling step was added for ultra-high coverage sequencing data using sambamba (0.7.1)^90^. For amplicon sequencing, assembly was conducted using pilon (1.24)^91^. Relevant analysis reports and visualization modules were also provided, including qualimap (2.2.2-dev)^92^ to analyze alignment metrics, snpEff (5.0e)^93^ to annotate SNPs, and QUAST (5.0.2)^94^ to inspect and visualize assembly results.

### Phylogenetic analysis

To elucidate the early phylogenetic relationships of SARS-CoV-2, we curated a dataset comprising 2,081 high-quality genomes and coverage gathered from the GISAID database up to March 1, 2020. To dissect the evolutionary trajectories of SARS-CoV-2 at genomic sites 8782 and 28144, our analysis augmented the 302-genome subset of Variants of Concern (VOCs) with a diverse selection of lineage-specific samples. This expansion included 600 samples from lineage A, 1,000 from lineage B, 250 from the haplotype 8782 C>T, and 80 from the 28144 T>C. We utilized the Nextstrain framework nextstrain (4.0.0)^95^ to construct the phylogenetic tree, anchoring the analysis with the Wuhan-Hu-1 strain as the root reference. The resulting phylogenetic tree was visualized using the R package ggtree (3.2.1)^96^.

To explore the evolutionary history of the genomic sites 8782 and 28144, we conducted a genomic alignment of 17 representative SARS-CoV-2 viruses and 40 other highly related viruses, including RpYN06, PrC31, and BANALs that were reported in 2021 and 2022^97–99^. The genomic sequences were aligned using MAFFT with the option “--auto”. We then used iqtree (2.0.3)^100^ with options “-bb 1000 -alrt 1000 -nt 64 -asr” to construct a 1000-times bootstrapped maximum-likelihood phylogenetic tree of the 57 viral sequences. The resulting phylogenetic tree was visualized using ggtreeExtra (1.4.2)^101^.

### Lineage assignment

The GISAID nomenclature (https://gisaid.org/resources/statements-clarifications/clade-and-lineage-nomenclature-aids-in-genomic-epidemiology-of-active-hcov-19-viruses/) shown in the phylogenetic tree was assigned based on the phylogeny and mutations noted using nextstrain (4.0.0). The SARS-CoV-2 started by lineages A and B (or S and L) based on the mutation L84S in the ORF8 protein^7^, denoted as 19A and 19B. We further divided them into four variants: lineage A (C8782T and T28144C), lineage B (C8782 and T28144), 8782 C>T (C8782T and T28144), and 28144 T>C (C8782 and T28144C). The GISAID clades were further refined with more detailed lineages assigned using the Phylogenetic Assignment of Named Global Outbreak LINeages tool, pangolin (4.1.3)^102^.

### SNP extraction from the GISAID database

The 5.5 million sequences from the GISAID database were aligned using MAFFT (7.505)^103^ with the “--auto” option. SNPs were then extracted using a custom Python script.

### Global distribution of lineages A and B

The regional information was extracted from the metadata of the GISAID database, with cities manually merged into provinces in China. The longitudes and latitudes of the areas were obtained through manual investigation using OpenStreetMap (http://api.openstreetmap.org/).

### Growth rate estimation

The growth rate r was calculated from the simulated daily case counts *π*(t) between t_1_ and t_2_ using the equation: r = log(*π*(t_2_)/*π*(t_1_))/(t_2_-t_1_). This formula represents the natural logarithm of the ratio of consecutive day counts. To address numerical instability from low case counts, days with counts below a threshold of one were excluded from the calculation to prevent unrealistic growth rate estimations.

### Reproduction Number

In the SIR (Susceptible-Infectious-Recovered) model of epidemiology, the reproduction number is presented as R_0_ = β/*γ*^57^, where β is the transmission rate per contact per day, and *γ* is the removal rate per day.

### The evoSIR model

The evoSIR model incorporates both the intra- and inter-host modeling, based on the classical Susceptible-Infected-Removed (SIR) model, to simulate the competitive dynamic of infectious diseases.

The model intra-host incorporates terms for the replication rate, competition during co-infection, and viral evolution. The intra-host model of multiple competing variants 1, 2, …, J follows the ordinary differential equations (ODEs):

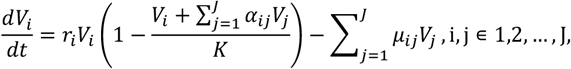

where V_i_ and V_j_ are the titers of variants i and j, and r_i_ is the replication rate of variant i. a_ij_ represents the inhibitory effect of variant j on variant i, and μ_ij_ represents the mutation rate of the virus from variant j to variant i.

For the inter-host component, we extended the SIR model to include compartments for individuals infected with each variant. S is the number of susceptible individuals correlated with the spreading capacity Q, which represents the susceptible contacts per infected individual; I_j_ is the individuals infected by variant j, and R is the number of removed individuals. N is the total population (N=S+I_1_+I_2_+R). The inter-host model follows:

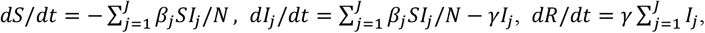

where 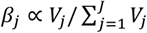 is the transmission rate for variant j, *γ* is the removal (recovery or death) rate for infected individuals. The expected number of new cases for variant j is β_j_SI/N per day.

### Estimation of parameters in the evoSIR model

For intra-host dynamics, parameters were estimated using experimental data scaled relative to the minimum titer in the samples, applying maximum likelihood estimation (MLE) (Table S1). Initial conditions were set as V = 1 unit, and the model was evaluated over a 72-unit period with a time step of 0.1 unit. We assume exponential growth in the initial period to simplify the ODEs as first-order linear differential equations: dV/dt = rV. The solution depends on the exponential growth of r_i_: V_t_ = V_0_e^rt^. The carrying capacity K was estimated from the maximum change of the viral titers. The system stability was assessed by calculating the Jacobian matrix at equilibrium points using rootSolve and number packages.

For inter-host dynamics, we estimated the parameters to fit the daily incidence data of COVID-19 in Wuhan from January 1 to March 8, 2020^23^. We specified the initial state of the model based on December 29 to December 31, 2019 (Table S2). We estimated β and *γ* by Markov chain Monte Carlo (MCMC) using rstan (2.21.8) (https://mc-stan.org/). We used an initial prior of Normal(0.3,0.1) for β and Normal(0.1,0.05) for *γ*. The likelihood for MCMC fitting was calculated using a Poisson distribution. A constraint maintained the A-to-B case ratio at 3:7, using a normal distribution centered at 0.3 with a standard deviation of 0.005. We set a burn-in period of 10,000 iterations and continued to run 15,000 iterations. Four chains were run, and the performance of the model fitting was checked by looking at measures of the bulk-ESS (greater than 100), tail-ESS (greater than 100), and individual parameters’ potential scale reduction statistic, 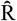 (less than 1.05)^65^. Trace plots for the model parameters were also generated.

### Sensitivity analysis

We conducted a sensitivity analysis by varying the initial frequency of lineage A from 0.1 to 0.9 in increments of 0.1 (Table S3). The mean value and 95% credible interval were shown. Assuming that the virus transmits in the early exponential growth stage, the difference between the replication rate was calculated as r_2_ − r_1_ = −log(β_1_/β_2_). The difference between the introduction dates of the two variants was calculated as log(I_1_)/(β_1_-*γ*) − log(I_2_)/(β_2_-*γ*).

### The evoSIR-Seeding-Spreading (evoSSS) model

The evoSIR-Seeding-Spreading (evoSSS) model integrated a Seeding-Spreading framework with the evoSIR model. The transition from one cycle to the next is represented by the composition of the seeding process and the evoSIR model:

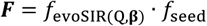

This composition function is applied to the matrix of infectious individuals, transforming the state from cycle n to n+1:

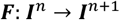

where **I**^n^ is the matrix representing the time sequence of infectious individuals for variants 1,2,…,J. The seeding process generates infectious seeds **D**^n^=[D_1_,D_2_,…,D_J_] from infectious individuals **I**^n^=[I_1_,I_2_,…,I_J_],

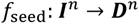

utilizes a mobility matrix 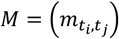 K to simulate the generation of epidemic hotspots from the infectious seeds at different time points. The elements of the mobility matrix are defined as

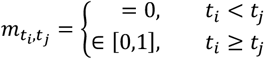

This process generates infectious seeds

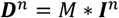

These infectious seeds create epidemic hotspots at each time point. Each hotspot h undergoes the evoSIR modeling

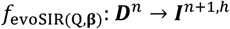

where the spreading capacity Q determines the susceptible individuals per seed, while transmission rates **β**=[β_1_,β_2,…,_β_J_] indicate the variant risk. The epidemic outcomes of these hotspots are then aggregated to represent the global pandemic state for the next epidemic cycle:

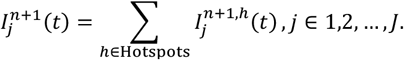

Specifically, the simulation in this paper used 200 days and an interspace of 30 days for each seeding-spreading cycle.

### Estimations of parameters in the evoSSS model

The fitting of the evoSSS model includes the estimations of mobility, spread capacity, and transmission rates for each variant. The initial state was specified by the observed data before the estimation period. The fitting utilized a self-adapting approach along with the seeding-spreading cycle by MCMC with rstan (2.21.8) (https://mc-stan.org/). The initial priors for the spreading capacity and transmission rate were set as Normal(200, 10) and Normal(0.3, 0.01), respectively. The removal rate (*γ*) was set as 0.157 according to the Wuhan fitting results. For successive cycles, the priors were drawn from a normal distribution based on the previous fitting results. We applied a scaling factor of (1e-5 * the maximum order of magnitude) to each simulation of initial infectious individuals to ensure that the variant could persist due to random mutations, which prevented biased estimations of transmission rates. The expected values of onset infectious individuals for the next spreading cycle were calculated as the observed infectious individuals minus the simulated onset infectious individuals from the previous cycle over 60 days, followed by a zero constraint applied for the next 40 days. The likelihood of observing the data given the model parameters was modeled using a Poisson distribution. We set a burn-in period of 2,000 iterations and continued to run 3,000 iterations. Four chains were run, and the performance of the model fitting was checked by looking at measures of the bulk-ESS (greater than 100), tail-ESS (greater than 100), and individual parameters’ potential scale reduction statistic, 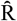 (less than 1.05)^65^. The goodness of fit is further measured by the Kolmogorov-Smirnov (KS) statistic with p-value^104^.

### Modeling and stochastic simulations

Euler’s numerical modeling was implemented, with multinomial sampling as stochastic simulations.

Intra-host dynamics models the competition between viral strains at the individual level. Stochastic effects are incorporated by sampling changes in the viral titer states of V_j_ using a multinomial distribution. The total change rate is calculated as the sum of the growth rates of both strains total_change_rate = ∑dV_j_/dt. The probability of change for each viral strain is determined by normalizing their respective growth rates: p_j_ = dV_j_/dt / total_change_rate. Changes in population are sampled with the probabilities p_Vj_:

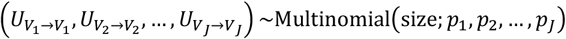

where size = ⌊total_change_rate⌋ + as.numeric(runif(1) < (total_change_rate − ⌊total_change_rate⌋)).

For inter-host modeling, we first generated seeds using a multinomial distribution

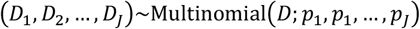

where D is the total number of seed virions or seed populations, and p_j_ represents the proportion of each variant. Then, the spreading process is implemented using a series of multinomial transitions. The transition from susceptible to infected with variant j follows:

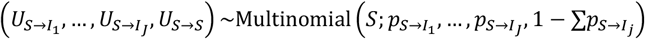

And the transition from infected to removed follows:

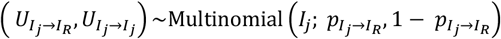

The transition probabilities are defined as p_S→Ij_ = β_j_S/N and p_Ij→R_ = *γ*.

## Data Availability

All data produced in the present work are contained in the manuscript

## Acknowledgments

We thank Xiangwei He for assisting with the paper revision. We extend our gratitude to our colleagues at the core facility of the Life Sciences Institute, particularly the NECHO high-performance computing cluster. We also gratefully acknowledge the authors from the originating and submitting laboratories, who generated and shared via GISAID genetic sequence data on which this research is based. This research was supported by grants from Key Research and Development Program of Zhejiang Province (2021C03039) and was partly supported by grants from National Natural Science Foundation of China NSFC (82173645, 82341109 and U20A20343) and Fundamental Research Funds for the Central Universities (2022ZFJH003).

## Author contributions

C.J. conceived the study, secured funding, and provided overall supervision. M.Z. and H.Y. designed the strategy for collecting clinical data and samples. S.H. contributed to the methodology and conducted formal analysis and visualization. X.L. and H.Y. carried out the experiments, while S.H., X.W., and Z.F curated the epidemiological data. The original draft was written by S.H., with review and editing done by C.J. and M.L. Z.X., Y.Z., L.H., and S.Z. provided suggestions for revising the manuscript. All authors critically reviewed and approved the final manuscript.

## Competing interests

The authors declare no competing interests.

## Data availability

The NGS sequencing data have been deposited to the NCBI Sequence Read Archive database (https://www.ncbi.nlm.nih.gov/sra) under BioProject PRJNA1153697, with relevant information included as Data S1. A total of 5,517,663 submissions in the GISAID database are included in this study. A complete list of 5.5 million accession numbers is included as Data S2.

## Code availability

The source code for the data processing and analyses is available at https://github.com/Hc1023/EvoSSS.

## Notes

### Competing Interest Statement

The authors have declared no competing interest.

### Author Declarations

The Ethics Committee of The First Affiliated Hospital, School of Medicine, Zhejiang University gave ethical approval for this work (Approval notice 2020-29) for emerging infectious diseases.

## References

1. Foraker, R. E. et al. Transmission dynamics: data sharing in the COVID-19 era. Learning Health Systems 5, e10235 (2021).

2. Rambaut, A. et al. A dynamic nomenclature proposal for SARS-CoV-2 lineages to assist genomic epidemiology. Nat Microbiol 5, 1403–1407, doi:10.1038/s41564-020-0770-5 (2020).

3. Tegally, H. et al., doi:10.1101/2020.12.21.20248640 (2020).

4. Ong, S. W. X. et al. Clinical and virological features of severe acute respiratory syndrome coronavirus 2 (SARS-CoV-2) variants of concern: a retrospective cohort study comparing B. 1.1. 7 (Alpha), B. 1.351 (Beta), and B. 1.617. 2 (Delta). Clinical Infectious Diseases 75, e1128–e1136 (2022).

5. Choi, J. Y. & Smith, D. M. SARS-CoV-2 Variants of Concern. Yonsei Med J 62, 961–968, doi:10.3349/ymj.2021.62.11.961 (2021).

6. Peacock, T. P. et al. The SARS-CoV-2 variant, Omicron, shows rapid replication in human primary nasal epithelial cultures and efficiently uses the endosomal route of entry. BioRxiv, 2021.2012. 2031.474653 (2022).

7. Forster, P., Forster, L., Renfrew, C. & Forster, M. Phylogenetic network analysis of SARS-CoV-2 genomes. Proc Natl Acad Sci U S A 117, 9241–9243, doi:10.1073/pnas.2004999117 (2020).

8. Tang, X. et al. On the origin and continuing evolution of SARS-CoV-2. Natl Sci Rev 7, 1012–1023, doi:10.1093/nsr/nwaa036 (2020).

9. Lu, R. et al. Genomic characterisation and epidemiology of 2019 novel coronavirus: implications for virus origins and receptor binding. The lancet 395, 565–574 (2020).

10. Wu, F. et al. A new coronavirus associated with human respiratory disease in China. Nature 579, 265–269 (2020).

11. Lytras, S. et al. Exploring the natural origins of SARS-CoV-2 in the light of recombination. Genome Biology and Evolution 14, evac018 (2022).

12. Holmes, E. C. et al. The origins of SARS-CoV-2: A critical review. Cell 184, 4848–4856, doi:10.1016/j.cell.2021.08.017 (2021).

13. de Sousa, E. et al. Mortality in COVID-19 disease patients: Correlating the association of major histocompatibility complex (MHC) with severe acute respiratory syndrome 2 (SARS-CoV-2) variants. Int J Infect Dis 98, 454–459, doi:10.1016/j.ijid.2020.07.016 (2020).

14. Ohki, S., Imamura, T., Higashimura, Y., Matsumoto, K. & Mori, M. Similarities and differences in the conformational stability and reversibility of ORF8, an accessory protein of SARS-CoV-2, and its L84S variant. Biochem Biophys Res Commun 563, 92–97, doi:10.1016/j.bbrc.2021.05.074 (2021).

15. Aoki, A., Mori, Y., Okamoto, Y. & Jinno, H. Development of a genotyping platform for SARS-CoV-2 variants using high-resolution melting analysis. J Infect Chemother 27, 1336–1341, doi:10.1016/j.jiac.2021.06.007 (2021).

16. Flower, T. G. et al. Structure of SARS-CoV-2 ORF8, a rapidly evolving immune evasion protein. Proceedings of the National Academy of Sciences 118, e2021785118 (2021).

17. Garry, R. F. Early appearance of two distinct genomic lineages of SARS-CoV-2 in different wuhan wildlife markets suggests SARS-CoV-2 has a natural origin. Virological 900, 110 (2021).

18. Pekar, J. E. et al. The molecular epidemiology of multiple zoonotic origins of SARS-CoV-2. Science 377, 960–966 (2022).

19. Blueprint, R. List of Blueprint priority diseases. World Health Organization [Internet] (2018).

20. Tahir, M. J. et al. Disease X: A hidden but inevitable creeping danger. Infection Control & Hospital Epidemiology 43, 1758–1759 (2022).

21. Anderson, R. M. & May, R. M. Population biology of infectious diseases: Part I. Nature 280, 361–367 (1979).

22. May, R. M. & Anderson, R. M. Population biology of infectious diseases: Part II. Nature 280, 455–461 (1979).

23. Hao, X. et al. Reconstruction of the full transmission dynamics of COVID-19 in Wuhan. Nature 584, 420–424, doi:10.1038/s41586-020-2554-8 (2020).

24. Anastassopoulou, C., Russo, L., Tsakris, A. & Siettos, C. Data-based analysis, modelling and forecasting of the COVID-19 outbreak. PLoS One 15, e0230405, doi:10.1371/journal.pone.0230405 (2020).

25. Giordano, G. et al. Modelling the COVID-19 epidemic and implementation of population-wide interventions in Italy. Nat Med 26, 855–860, doi:10.1038/s41591-020-0883-7 (2020).

26. Kyrychko, Y. N., Blyuss, K. B. & Brovchenko, I. Mathematical modelling of the dynamics and containment of COVID-19 in Ukraine. Scientific reports 10, 19662 (2020).

27. Gatto, M. et al. Spread and dynamics of the COVID-19 epidemic in Italy: Effects of emergency containment measures. Proceedings of the National Academy of Sciences 117, 10484–10491 (2020).

28. Holmes, E. C. & Grenfell, B. T. Discovering the phylodynamics of RNA viruses. PLoS computational biology 5, e1000505 (2009).

29. Keeling, M. J. & Rohani, P. Modeling infectious diseases in humans and animals. (Princeton university press, 2011).

30. Ball, F. et al. Seven challenges for metapopulation models of epidemics, including households models. Epidemics 10, 63–67 (2015).

31. Grenfell, B. T., Bjørnstad, O. N. & Kappey, J. Travelling waves and spatial hierarchies in measles epidemics. Nature 414, 716–723 (2001).

32. Heesterbeek, H. et al. Modeling infectious disease dynamics in the complex landscape of global health. Science 347, aaa4339 (2015).

33. Anderson, R. M. & May, R. M. Infectious diseases of humans: dynamics and control. (Oxford university press, 1991).

34. Andersen, K. G., Rambaut, A., Lipkin, W. I., Holmes, E. C. & Garry, R. F. The proximal origin of SARS-CoV-2. Nat Med 26, 450–452, doi:10.1038/s41591-020-0820-9 (2020).

35. Pepin, K. M., Lass, S., Pulliam, J. R., Read, A. F. & Lloyd-Smith, J. O. Identifying genetic markers of adaptation for surveillance of viral host jumps. Nature Reviews Microbiology 8, 802–813 (2010).

36. Schluter, D. The ecology of adaptive radiation. (OUP Oxford, 2000).

37. Smith, V. H. & Holt, R. D. Resource competition and within-host disease dynamics. Trends in ecology & evolution 11, 386–389 (1996).

38. Pybus, O. G. & Rambaut, A. Evolutionary analysis of the dynamics of viral infectious disease. Nature Reviews Genetics 10, 540–550 (2009).

39. Grenfell, B. T. et al. Unifying the epidemiological and evolutionary dynamics of pathogens. Science 303, 327–332 (2004).

40. Kimura, M. The neutral theory of molecular evolution. (Cambridge University Press, 1983).

41. Yao, H. et al. Patient-derived SARS-CoV-2 mutations impact viral replication dynamics and infectivity in vitro and with clinical implications in vivo. Cell Discov 6, 76, doi:10.1038/s41421-020-00226-1 (2020).

42. Tonkin-Hill, G. et al. Patterns of within-host genetic diversity in SARS-CoV-2. Elife 10, doi:10.7554/eLife.66857 (2021).

43. Rockett, R. J. et al. Co-infection with SARS-CoV-2 Omicron and Delta variants revealed by 4 genomic surveillance. Nat Commun 13, 2745, doi:10.1038/s41467-022-30518-x (2022).

44. Wang, Y. et al. Intra-host variation and evolutionary dynamics of SARS-CoV-2 populations in COVID-19 patients. Genome Med 13, 30, doi:10.1186/s13073-021-00847-5 (2021).

45. Ulrich, L. et al. Enhanced fitness of SARS-CoV-2 variant of concern Alpha but not Beta. Nature 602, 307–313, doi:10.1038/s41586-021-04342-0 (2022).

46. Plante, J. A. et al. Spike mutation D614G alters SARS-CoV-2 fitness. Nature 592, 116–121, doi:10.1038/s41586-020-2895-3 (2021).

47. Liu, Y. et al. Delta spike P681R mutation enhances SARS-CoV-2 fitness over Alpha variant. Cell Rep 39, 110829, doi:10.1016/j.celrep.2022.110829 (2022).

48. Markov, P. V. et al. The evolution of SARS-CoV-2. Nat Rev Microbiol 21, 361–379, doi:10.1038/s41579-023-00878-2 (2023).

49. Pan, A. et al. Association of public health interventions with the epidemiology of the COVID-19 outbreak in Wuhan, China. Jama 323, 1915–1923 (2020).

50. Miller, J. K., Elenberg, K. & Dubrawski, A. Forecasting emergence of COVID-19 variants of concern. PLoS One 17, e0264198 (2022).

51. Fitzpatrick, M. C., Bauch, C. T., Townsend, J. P. & Galvani, A. P. Modelling microbial infection to address global health challenges. Nature microbiology 4, 1612–1619 (2019).

52. Wiratsudakul, A., Suparit, P. & Modchang, C. Dynamics of Zika virus outbreaks: an overview of mathematical modeling approaches. PeerJ 6, e4526 (2018).

53. Ignatov, A. & Trigger, S. Two viruses competition in the SIR model of epidemic spread: application to COVID-19. MedRxiv, 2022.2001. 2011.22269046 (2022).

54. Zhang, C., Gracy, S., BaŞar, T. & Paré, P. E. A networked competitive multi-virus SIR model: Analysis and observability. IFAC-PapersOnLine 55, 13–18 (2022).

55. GF, G. Experimental studies on the struggle for existence. J Exp Biol 9, 389–402 (1932).

56. Grover, J. P. Resource competition. Vol. 19 (Springer Science & Business Media, 1997).

57. Kermack, W. O. & McKendrick, A. G. A contribution to the mathematical theory of epidemics. Proceedings of the royal society of london. Series A, Containing papers of a mathematical and physical character 115, 700–721 (1927).

58. Zhang, X. et al. Viral and host factors related to the clinical outcome of COVID-19. Nature 583, 437–440, doi:10.1038/s41586-020-2355-0 (2020).

59. Pawitan, Y. In all likelihood: statistical modelling and inference using likelihood. (Oxford University Press, 2001).

60. Brooks, S. Markov chain Monte Carlo method and its application. Journal of the royal statistical society: series D (the Statistician) 47, 69–100 (1998).

61. Furukawa, N. W., Brooks, J. T. & Sobel, J. Evidence supporting transmission of severe acute respiratory syndrome coronavirus 2 while presymptomatic or asymptomatic. Emerging infectious diseases 26 (2020).

62. Díez-Fuertes, F. et al. A founder effect led early SARS-CoV-2 transmission in Spain. Journal of Virology 95, 10.1128/jvi. 01583-01520 (2021).

63. Farkas, C., Fuentes-Villalobos, F., Garrido, J. L., Haigh, J. & Barría, M. I. Insights on early mutational events in SARS-CoV-2 virus reveal founder effects across geographical regions. PeerJ 8, e9255 (2020).

64. Wright, S. Evolution in Mendelian populations. Genetics 16, 97 (1931).

65. Vehtari, A., Gelman, A., Simpson, D., Carpenter, B. & Bürkner, P.-C. Rank-normalization, folding, and localization: An improved R ^ for assessing convergence of MCMC (with discussion). Bayesian analysis 16, 667–718 (2021).

66. Volz, E. M., Koelle, K. & Bedford, T. Viral phylodynamics. PLoS computational biology 9, e1002947 (2013).

67. Flahault, A. et al. FluNet as a tool for global monitoring of influenza on the Web. Jama 280, 1330–1332 (1998).

68. Giacinti, J. A. et al. Avian influenza viruses in wild birds in Canada following incursions of highly pathogenic H5N1 virus from Eurasia in 2021–2022. mBio, e03203-03223 (2024).

69. Kojima, N. et al. Building global preparedness for avian influenza. The Lancet 403, 2461–2465 (2024).

70. Ulaeto, D. et al. New nomenclature for mpox (monkeypox) and monkeypox virus clades. The Lancet Infectious Diseases 23, 273–275 (2023).

71. Schuele, L. et al. Real-time PCR assay to detect the novel Clade Ib monkeypox virus, September 2023 to May 2024. Eurosurveillance 29, 2400486 (2024).

72. Zheng, L. et al. Global variability of influenza activity and virus subtype circulation from 2011 to 2023. BMJ Open Respiratory Research 10, e001638 (2023).

73. Zeng, Z. et al. Molecular epidemiology and phylogenetic analysis of influenza viruses A (H3N2) and B/Victoria during the COVID-19 pandemic in Guangdong, China. Infectious Diseases of Poverty 13, 56 (2024).

74. Rohani, P. & Bahl, J. Collateral effects of pandemic control. Science 386, 620–621 (2024).

75. Chen, Z. et al. COVID-19 pandemic interventions reshaped the global dispersal of seasonal influenza viruses. Science 386, eadq3003 (2024).

76. Barut, G. T. et al. The spike gene is a major determinant for the SARS-CoV-2 Omicron-BA.1 phenotype. Nat Commun 13, 5929, doi:10.1038/s41467-022-33632-y (2022).

77. Nagy, A., Pongor, S. & Gyorffy, B. Different mutations in SARS-CoV-2 associate with severe and mild outcome. Int J Antimicrob Agents 57, 106272, doi:10.1016/j.ijantimicag.2020.106272 (2021).

78. Bendall, E. E. et al. SARS-CoV-2 genomic diversity in households highlights the challenges of sequence-based transmission inference. Msphere 7, e00400–00422 (2022).

79. Memish, Z. A. et al. Proactive surveillance for avian influenza H5N1 and other priority pathogens at mass gathering events. The Lancet Public Health 9, e350–e352 (2024).

80. Yurkovetskiy, L. et al. Structural and Functional Analysis of the D614G SARS-CoV-2 Spike Protein Variant. Cell 183, 739–751 e738, doi:10.1016/j.cell.2020.09.032 (2020).

81. Zhang, J. et al. Membrane fusion and immune evasion by the spike protein of SARS-CoV-2 Delta variant. Science 374, 1353–1360 (2021).

82. Roda, W. C., Varughese, M. B., Han, D. & Li, M. Y. Why is it difficult to accurately predict the COVID-19 epidemic? Infectious Disease Modelling 5, 271–281, doi:10.1016/j.idm.2020.03.001 (2020).

83. Chen, S., Zhou, Y., Chen, Y. & Gu, J. fastp: an ultra-fast all-in-one FASTQ preprocessor. Bioinformatics 34, i884–i890 (2018).

84. Langmead, B. & Salzberg, S. L. Fast gapped-read alignment with Bowtie 2. Nature methods 9, 357–359 (2012).

85. Li, H. et al. The sequence alignment/map format and SAMtools. Bioinformatics 25, 2078–2079 (2009).

86. Au, C. H., Ho, D. N., Kwong, A., Chan, T. L. & Ma, E. S. K. BAMClipper: removing primers from alignments to minimize false-negative mutations in amplicon next-generation sequencing. Sci Rep 7, 1567, doi:10.1038/s41598-017-01703-6 (2017).

87. Koboldt, D. C. et al. VarScan 2: somatic mutation and copy number alteration discovery in cancer by exome sequencing. Genome Res 22, 568–576, doi:10.1101/gr.129684.111 (2012).

88. Li, H. Aligning sequence reads, clone sequences and assembly contigs with BWA-MEM. arXiv preprint 1303.3997 (2013).

89. Bankevich, A. et al. SPAdes: a new genome assembly algorithm and its applications to single-cell sequencing. Journal of computational biology 19, 455–477 (2012).

90. Tarasov, A., Vilella, A. J., Cuppen, E., Nijman, I. J. & Prins, P. Sambamba: fast processing of NGS alignment formats. Bioinformatics 31, 2032–2034 (2015).

91. Walker, B. J. et al. Pilon: an integrated tool for comprehensive microbial variant detection and genome assembly improvement. PloS one 9, e112963 (2014).

92. Okonechnikov, K., Conesa, A. & García-Alcalde, F. Qualimap 2: advanced multi-sample quality control for high-throughput sequencing data. Bioinformatics 32, 292–294 (2016).

93. Cingolani, P. et al. A program for annotating and predicting the effects of single nucleotide polymorphisms, SnpEff: SNPs in the genome of Drosophila melanogaster strain w1118; iso-2; iso-3. Fly 6, 80–92 (2012).

94. Gurevich, A., Saveliev, V., Vyahhi, N. & Tesler, G. QUAST: quality assessment tool for genome assemblies. Bioinformatics 29, 1072–1075 (2013).

95. Hadfield, J. et al. Nextstrain: real-time tracking of pathogen evolution. Bioinformatics 34, 4121–4123, doi:10.1093/bioinformatics/bty407 (2018).

96. Yu, G., Smith, D. K., Zhu, H., Guan, Y. & Lam, T. T. Y. ggtree: an R package for visualization and annotation of phylogenetic trees with their covariates and other associated data. Methods in Ecology and Evolution 8, 28–36 (2017).

97. Zhou, H. et al. Identification of novel bat coronaviruses sheds light on the evolutionary origins of SARS-CoV-2 and related viruses. Cell 184, 4380–4391 e4314, doi:10.1016/j.cell.2021.06.008 (2021).

98. Li, L. L. et al. A novel SARS-CoV-2 related coronavirus with complex recombination isolated from bats in Yunnan province, China. Emerg Microbes Infect 10, 1683–1690, doi:10.1080/22221751.2021.1964925 (2021).

99. Temmam, S. et al. Bat coronaviruses related to SARS-CoV-2 and infectious for human cells. Nature 604, 330–336, doi:10.1038/s41586-022-04532-4 (2022).

100. Minh, B. Q. et al. IQ-TREE 2: new models and efficient methods for phylogenetic inference in the genomic era. Molecular biology and evolution 37, 1530–1534 (2020).

101. Xu, S. et al. ggtreeExtra: Compact visualization of richly annotated phylogenetic data. Molecular biology and evolution 38, 4039–4042 (2021).

102. O’Toole, Á. et al. Assignment of epidemiological lineages in an emerging pandemic using the pangolin tool. Virus evolution 7, veab064 (2021).

103. Katoh, K. & Standley, D. M. MAFFT multiple sequence alignment software version 7: improvements in performance and usability. Molecular biology and evolution 30, 772–780 (2013).

104. An, K. Sulla determinazione empirica di una legge didistribuzione. Giorn Dell’inst Ital Degli Att 4, 89–91 (1933).

